# Screening for Maternally Inherited Diabetes and Deafness in Large Cohorts of Hearing Impaired and Diabetic Patients

**DOI:** 10.1101/2025.03.13.25321027

**Authors:** Lukas Varga, Silvia Borecka, Martina Skopkova, Vibhuti Rambani, Marek Sklenar, Klaudia Cipkova, Terezia Kickova, Diana Ugorova, Zuzana Kabatova, Juraj Stanik, Milan Profant, Daniela Gasperikova

## Abstract

**Objectives:** Mitochondrial DNA (mtDNA) mutations account for up to 5% of hereditary hearing loss cases. Most commonly, the m.3243A>G mtDNA variant contributes to rare monogenic MIDD (Maternally Inherited Diabetes and Deafness) or MELAS (Mitochondrial Encephalopathy, Lactic Acidosis, and Stroke-like episodes) syndromes. Different proportions of the mutated mtDNA (heteroplasmy) among the affected tissues result in variability in the clinical manifestation and severity of the phenotype. **The aim** of the presented study was to establish the prevalence of the m.3243A>G variant in large cohorts of hearing-impaired and diabetic patients in Slovakia and to evaluate the genotype-phenotype correlations and long-term cochlear implantation outcomes.

**Design:** Probands (n=5957) were recruited via three independent nationwide studies on hereditary hearing loss (n=1145) and diabetes (unselected diabetes group, n=4158 and Monogenic diabetes group, n=654; total n=4812). DNA from peripheral blood and/or buccal mucosa was tested for the presence of the m.3243A>G variant using two PCR methods – qPCR and dPCR. Audiological and other clinical data of the identified variant carriers were also collected for phenotype evaluation.

**Results:** We identified 25 probands/families harboring the m.3243A>G variant (0.42%). The prevalence was higher in the groups where monogenic disorder was suspected – 0.79% in the Hearing loss group and 1.68% in the Monogenic diabetes group versus 0.14% in the general diabetes group (*p* < 0.001). Heteroplasmy levels assessed by dPCR ranged between 0.04% and 76% in peripheral blood and 0.01% and 92% in buccal samples. In most individuals, the symptoms manifested in the fourth decade of life in affected subjects with the MIDD phenotype or isolated hearing loss/diabetes, but as early as in the second decade in the probands with MELAS. We observed high phenotype variability, ranging from severe multisystemic involvement through isolated symptoms to asymptomatic young “dormant” or very low heteroplasmy carriers. Only 54% of individuals with the m.3243A>G variant had both diabetes and hearing loss. The heteroplasmy levels from buccal swabs showed a better correlation with the age of onset of both hearing loss and diabetes than the age-adjusted blood heteroplasmy. On the other hand, the age-adjusted blood heteroplasmy was associated with overall severity of the disease (i.e., with a higher number of clinical symptoms). We show that the most typical audiogram configurations are flat and sloping. Three individuals identified as cochlear implant recipients showed excellent and long-term stable functional outcomes. In addition, the authors report the first case of successful stapes surgery in a patient with confirmed mitochondrial disorder.

**Conclusions:** The diagnostic yield was higher in the deafness and monogenic diabetes groups than in the unselected diabetes group. Implementation of rigorous inclusion criteria requiring the presence of both diabetes and hearing loss may lead to a lower detection rate due to different or incomplete phenotype manifestation. Age-adjusted blood heteroplasmy levels seem to be a good predictor of overall severity of m.3243A>G-associated diseases, but buccal mucosa heteroplasmy better predicted the age of hearing loss and diabetes onset. We further confirm that cochlear implantation and stapedectomy are safe and efficient options for hearing restoration and rehabilitation in m.3243A>G carriers.

## Introduction

Both hearing loss (HL) and diabetes belong among the most prevalent chronic disorders at the global level, with hearing loss affecting one in five people (GBD 2021) and diabetes over 10.5% of the world’s adult population (Sun et al. 2022). The etiology of these two diseases is very heterogeneous and includes many different genetic and non-genetic factors. An increase in knowledge on monogenic forms of diabetes allowed to separate this diabetes subgroup from other diabetes types, such as type 1 (T1D) or type 2 diabetes (T2D). Monogenic diabetes accounts for approximately 1–5% of all diabetes cases, with more than 40 different genes involved (Hattersley et al. 2018; Zhang et al. 2021; American Diabetes Association Professional Practice Committee 2022; Greeley et al. 2022). The genetic background of sensorineural hearing loss is even more notable, counting more than 150 genes associated with nonsyndromic sensorineural hearing loss and no less than 400 different syndromic forms (Toriello & Smith 2013; Walls et al, 2024). In some cases, diabetes and hearing loss are caused by the same causal genetic factor, like in Wolfram syndrome (*WFS1* gene), Kearns-Sayre syndrome (large-scale mitochondrial DNA deletions), thiamine-responsive megaloblastic anemia (*SLC19A2*) and maternally inherited diabetes and deafness (*MT-TL1*, *MT-TK*, *MT-TE*) (Gruber & Pinhas-Hamiel 2022).

Mitochondrial DNA (mtDNA) variants account for about 5% of postlingual hereditary hearing loss cases in the Caucasian population (Jacobs et al. 2005). Certain pathogenic mtDNA variants also contribute to the rare monogenic form of diabetes (MIDD – Maternally Inherited Diabetes and Deafness, MIM 520000) or neurological disease (MELAS – Mitochondrial Myopathy, Encephalopathy, Lactic Acidosis, and Stroke-like episodes, MIM 540000). The most frequent variant causing these conditions is m.3243A>G (Finsterer & Frank 2016). Different proportions of the mutated mtDNA to wild-type mtDNA (heteroplasmy) among the affected tissues result in variability in the clinical manifestation and severity of the phenotype (Robinson et al. 2020). Heteroplasmy levels in peripheral blood leukocytes, in particular, are closely associated with clinical manifestations. They are valuable for the assessment of the clinical severity of the m.3243A>G variant in the affected individuals (Grady et al. 2018; Geng et al. 2019).

The m.3243A>G variant itself has a broad spectrum of clinical manifestations, including MIDD, MELAS, myoclonic epilepsy and ragged-red fibers (MERRF, MIM 545000), Kearns-Sayre syndrome (MIM 530000) or Leigh syndrome (MIM 256000). In many cases the variant carriers are oligosymptomatic or present with overlapping phenotypes (Dvorakova et al. 2016; de Laat et al. 2019; Li et al. 2022).

The presented study aimed to establish the prevalence of the m.3243A>G variant in large cohorts of hearing-impaired and diabetic patients in Slovakia. Additionally, we analyzed the phenotypes of the affected subjects, including the relationship of the onset of symptoms and their severity with mutant mtDNA load. Furthermore, we looked at the hearing rehabilitation outcomes of those subjects who underwent cochlear implantation (CI) or other surgical procedures to restore hearing.

## Materials and Methods

### Study source population

Probands (n = 5957) were recruited via three independent nationwide studies on the genetics of hearing loss (HL group, n = 1145) and diabetes (Diaret group, n = 4158 and Monogenic diabetes group, n = 654; total n = 4812).

The **HL group** included 1145 subjects with permanent bilateral hearing loss, with age of onset <60 years. Individuals with clearly non-genetic causes (e. g. meningitis, temporal bone trauma) were excluded. The hearing loss was of sensorineural type, with a few exceptions of those syndromic hearing loss forms in which the conductive component of HL may also occur. These subjects were referred from ENT departments in Slovakia or recruited at boarding schools for hearing-disabled children.

The **Diaret group** included 4158 individuals with diabetes mellitus (DM) recruited in the framework of an epidemiological multicenter survey of adult patients with diabetes in Slovakia DIARET SK (NCT02232503). The recruitment process and the cohort structure have been described elsewhere (Ondrejkova et al. 2019). Briefly, consecutive adult patients in a pre-specified sequence (every 5^th^, 10^th^ and 15^th^ patient) were included in the study with the exclusion of gestational or secondary induced diabetes, cases with ketoacidosis or hyperosmolar coma, and alcohol abuse. The cohort represented 1% of all people with DM in Slovakia at the time of recruitment.

**The monogenic diabetes group** consisted of 654 probands with clinical suspicion of monogenic diabetes. They were actively searched for in diabetes outpatient clinics, pediatric endocrine clinics and geneticists throughout Slovakia between the years 2003 – 2022. The inclusion criteria were based on previously published clinical guidelines for monogenic diabetes (Ellard et al. 2008; Hattersley et al. 2009) and included: age of DM onset <35 years, absence of pancreatic auto-antibodies or type 2 diabetes (obesity and signs of insulin resistance), and 3-generational DM. However, as the stringent application of these criteria could miss some patients (Stanik et al. 2014), only patients with positive antibodies and evident T2D were excluded. In addition, 40 matrilineal family members of probands with the m.3243A>G variant were recruited for the study.

The study was conducted in accordance with the Declaration of Helsinki of the World Medical Association. All individuals or their legal representatives (in subjects under 18 years of age) signed an informed consent to genetic testing, and the study was approved by the Ethics committee of the Bratislava self-governing region, the Ethics Committees of the University Hospital in Bratislava, the National Institute of Children’s Diseases in Bratislava and the National Institute of Endocrinology and Diabetes in Lubochna. Patients consented to having their images and other clinical information reported in the journal.

### Sampling

Samples of 8 ml of venous blood were collected into plasma separation tubes (BD Vacutainer) in the Diaret study or PAXgene Blood DNA tubes (Qiagene, UK) in the monogenic DM and HL group, and/or buccal swabs were taken for isolation of DNA for the molecular-genetic analysis. Clinical data about the disease character in the proband and family relatives and genealogic history were filled into the questionnaire by the attending physician. DNA testing was also extended to the family members of probands.

### Heteroplasmy assessment

The m.3243A>G heteroplasmy was measured using primers MIDD-F: 5’CCACACCCACCCAAGAACAG3’ and MIDD-R: 5’AGGAATTGAACCTCCGACTGTAAAGTTT3’ and mutation-specific fluorescently labeled ZNA probes (MIDDprobe WT: FAM - 5’CCGGGC<u>T</u>CTGCCAT3’- BHQ-1 and MIDDprobe MT: Yakima Yellow – 5’CCGGGC<u>C</u>CTGCCAT3’ - BHQ-1). Two methods for detection were used – real-time PCR (qPCR) and digital PCR (dPCR). For qPCR, TaqMan Gene Expression Master Mix, 0.9 µM primers, and 0.25 µM probes were run on QuantStudio 5 real-time cycler (LifeTechnologies). The heteroplasmy was counted as 2^ΔΔCt^/(1+ 2^ΔΔCt^) *100 and expressed in percentage of the mutant allele. For dPCR, Probe PCR MasterMix, 0.8 µM primers and 0.4 µM probes were run on the QIAcuity digital PCR system (Qiagen) in 26K 24-well plates. The heteroplasmy was counted as a mutant fraction by QIAcuity software from partition clusters adjusted manually in the 2D scatterplot option.

Age-adjusted heteroplasmy was counted from heteroplasmy detected in blood using dPCR. The formula used was hetadj = hetage/0.977^(age+12)^, as described in Grady et al. (2018).

### Phenotype characterization and clinical evaluation of the m.3243A>G variant carriers

Clinical history, audiological, biochemical, imaging and other clinical data of the identified variant carriers were collected for phenotype evaluation. Because the probands originated from three independent studies which were focused on different phenotype features, and the material and data collection was also designed differently in these studies (as described above), the availability of certain clinical data varied among the subjects. Questionnaires from all three studies contained questions on the presence of diabetes and metabolic disorders. However, only two of the three questionnaires (studies on Hereditary HL and Monogenic diabetes) contained data on hearing loss. To reduce these differences, probands with available contacts who agreed to participate in additional clinical evaluation were subsequently investigated in collaboration with their primary care physicians, ENT specialists or diabetologists. The relevant data from medical history, selected biochemical parameters and individual heteroplasmy levels are listed in Supplementary Table S1 and are available upon request from the corresponding author. The phenotype score was calculated as the number of symptoms manifested in the patient: hearing loss, diabetes, encephalopathy and other clinical symptoms – one point for each symptom.

### Statistical analysis

Statistical analysis was done using the open-source R Statistical Software package (RDevelopment team, 2006). Data were checked for normality using the Shapiro-Wilk test with the rstatix (0.7.0) package. The correlation of dPCR and qPCR values was tested using the Pearson correlation with the *stats* (4.4.1) package for R. Association of phenotype features and heteroplasmy levels were tested using linear regression with the *stats* (4.4.1) and forward stepwise multiple linear regression analysis with the *olsrr* (0.6.0) package for R. Ordered ANOVA for testing differences in heteroplasmy in groups with an increasing number of symptoms (phenotype score) was done using the Jonckheere-Terpstra test with the *PMCMRplus* (1.9.10) package for R. The post-hoc Tukey test was used for multiple comparison of means using the *stats* (4.4.1) package for R. Plots were prepared using the *ggplot2* (3.3.6) package for R. Differences between two groups of binary data were calculated via the chi square test.

## Results

### Prevalence of the m.3243A>G variant

Overall, we identified 25 probands/families harboring the m.3243A>G variant (0.42%) among the 5957 analyzed DNA samples. The prevalence was higher in the groups where the monogenic disorder was suspected – 0.79% in the hearing loss group (9/1145) and 1.68% in the Monogenic diabetes group (11/654) versus 0.14% in the general diabetes (Diaret) group (6/4158) (*p* < 0.001). One proband with the MELAS phenotype (III:1, Fam 3) was recruited independently in two study groups (hearing loss and monogenic diabetes). Subsequently, we analyzed the m.3243A>G variant in 40 matrilineal family members of the 25 positive probands who were available for genetic testing. In two of these family members the variant was not detected in the available sample material. Thus, the variant was detected in a total of 63 subjects (25 probands and 38 matrilineal family members).

### Heteroplasmy levels

First, we compared the two PCR methods (qPCR and dPCR) available for detection of the m.3243A>G variant and found a strong correlation (r = 0.97, *p* < 0.001) between the heteroplasmy levels determined by each of the methods regardless of the DNA material source (blood and buccal) (Supplementary Figure S1 in the Supplemental Digital Content). However, ten samples originally identified as negative by qPCR showed positive dPCR results, with dPCR heteroplasmy levels below 2.3%, confirming the higher sensitivity of this method.

In the tested subjects, heteroplasmy levels analyzed using dPCR from peripheral blood ranged between 0.04% (subject III:3, FAM15) and 76% (subject IV:1, FAM1), while from buccal swabs between 0.01% (subject III:3, FAM15) and 92% (subject IV:1, FAM16). The age- adjusted blood heteroplasmy ranged from 0.1% (subject III:3, FAM15) to 135% (subject III:1, FAM3) (Supplementary Table S1 available upon request from the corresponding author). The age-adjusted values exceed 100% due to the formula used, which accounts for the continuous decrease of heteroplasmy that is observed with age (Grady et al., 2018).

### Phenotypes

Phenotype variability in the 63 positive individuals was observed in the age of onset of symptoms and similarly in the severity of clinical manifestation. We identified 21 individuals with MIDD and 3 probands with MELAS syndrome. The remaining 39 individuals had hearing loss without diabetes (n = 12) or diabetes without hearing loss (n = 10), and 17 individuals had other symptoms or were asymptomatic (Supplementary Table S1 available upon request from the corresponding author).

Among subjects with confirmed **hearing loss** (n = 36), its severity ranged from mild high-frequency HL to profound HL/deafness. It was associated with age-adjusted blood heteroplasmy levels (R^2^ = 0.16, *p* = 0.0087; Figure 1), while no association was seen with buccal heteroplasmy levels (R^2^ = 0, *p* = 0.8; data not shown).

**Fig. 1.**
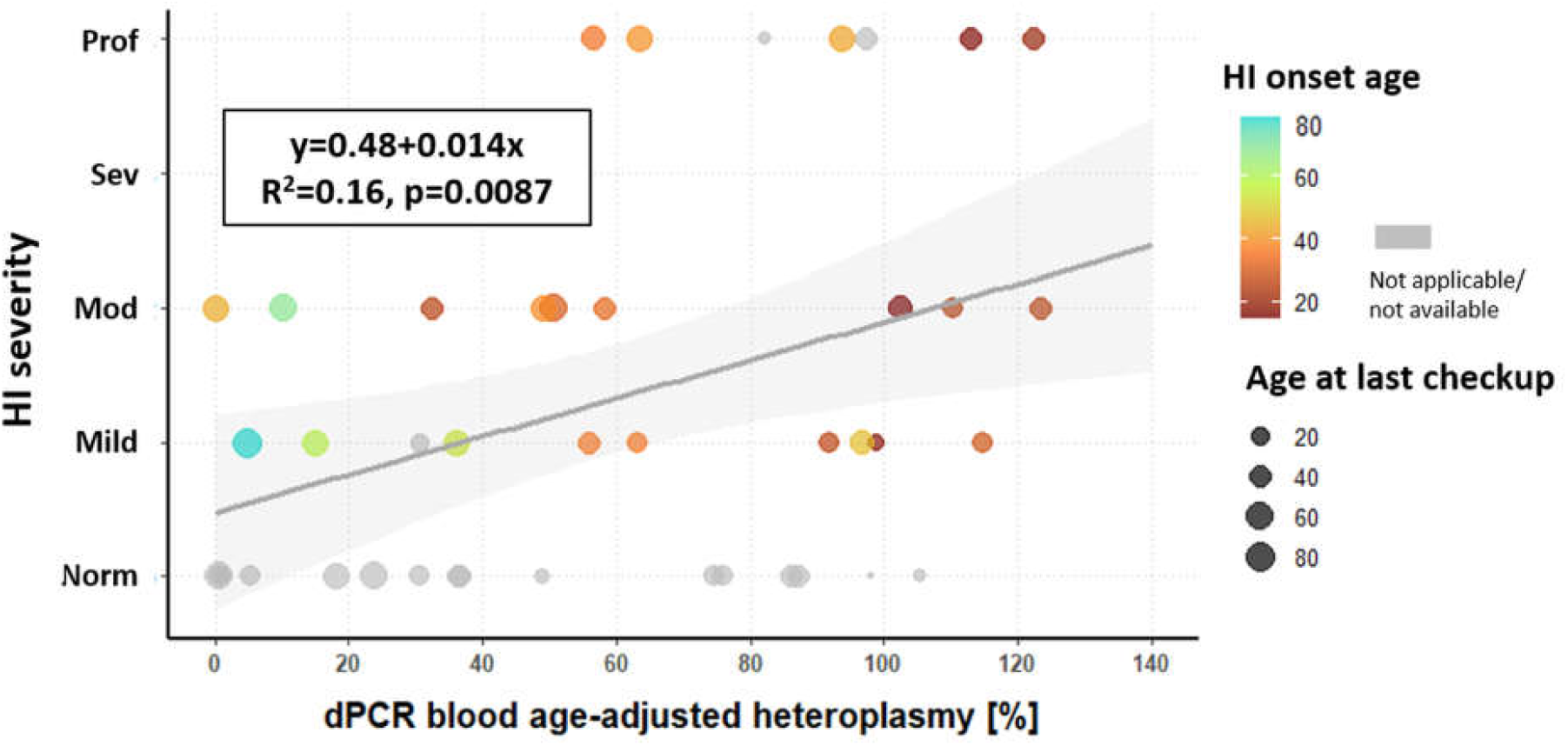
Association of severity of hearing loss with age-adjusted blood heteroplasmy of the m.3243G>A pathogenic variant. The color of the points expresses the age at HI onset, while the point size is relative to the age at the last checkup. dPCR – digital PCR; HI – hearing impairment.

We have also seen various audiogram configurations (Supplementary Figure S3 available upon request from the corresponding author). The most common audiogram shape was sloping with different degrees of steepness (n = 11), followed by flat (n = 10), residual hearing (n = 6), isolated high-frequency HL (n = 3), reverse cookie-bite audiogram (n = 1) and other (n=2, subject III:1 in FAM1 and subject III:2 in FAM 14) in Figure S3 available upon request from the corresponding author. Asymmetric HL was observed in 3 subjects. The HL was reported to be absent in 5 probands and 19 family members, although audiometry results were not provided for all of these subjects. Most of the subjects without HL were either of young age (10 subjects below the age of 20 years) or had low heteroplasmy levels (n = 9 with heteroplasmy <10%, of which in 2 subjects the variant was repeatedly below detection level). In 3 subjects (2 probands and 1 family member) the hearing status was not known from the available clinical data, and all were recruited in the diabetes branch of the study.

The onset of HL was reliably known in 29 subjects. Here, the average age of HL onset was 35.48 ± 16.78 years, ranging from 1 to 79 years (Supplementary Figure S2 in the Supplemental Digital Content). Linear regression analysis was performed to confirm the relationship between the age of hearing loss onset and the heteroplasmy levels. The heteroplasmy of buccal mucosa provides a better prediction of HL onset age (R^2^ = 0.66, *p* < 0.001, Figure 2A) than the age- adjusted blood heteroplasmy (R^2^ = 0.55, *p* < 0.001, Figure 2B). However, in a forward stepwise multiple linear regression analysis with both buccal mucosa and blood heteroplasmy as independent covariables, only buccal mucosa heteroplasmy was significantly associated with age of the hearing loss onset (β = -0.622, *p* < 0.001).

**Fig. 2.**
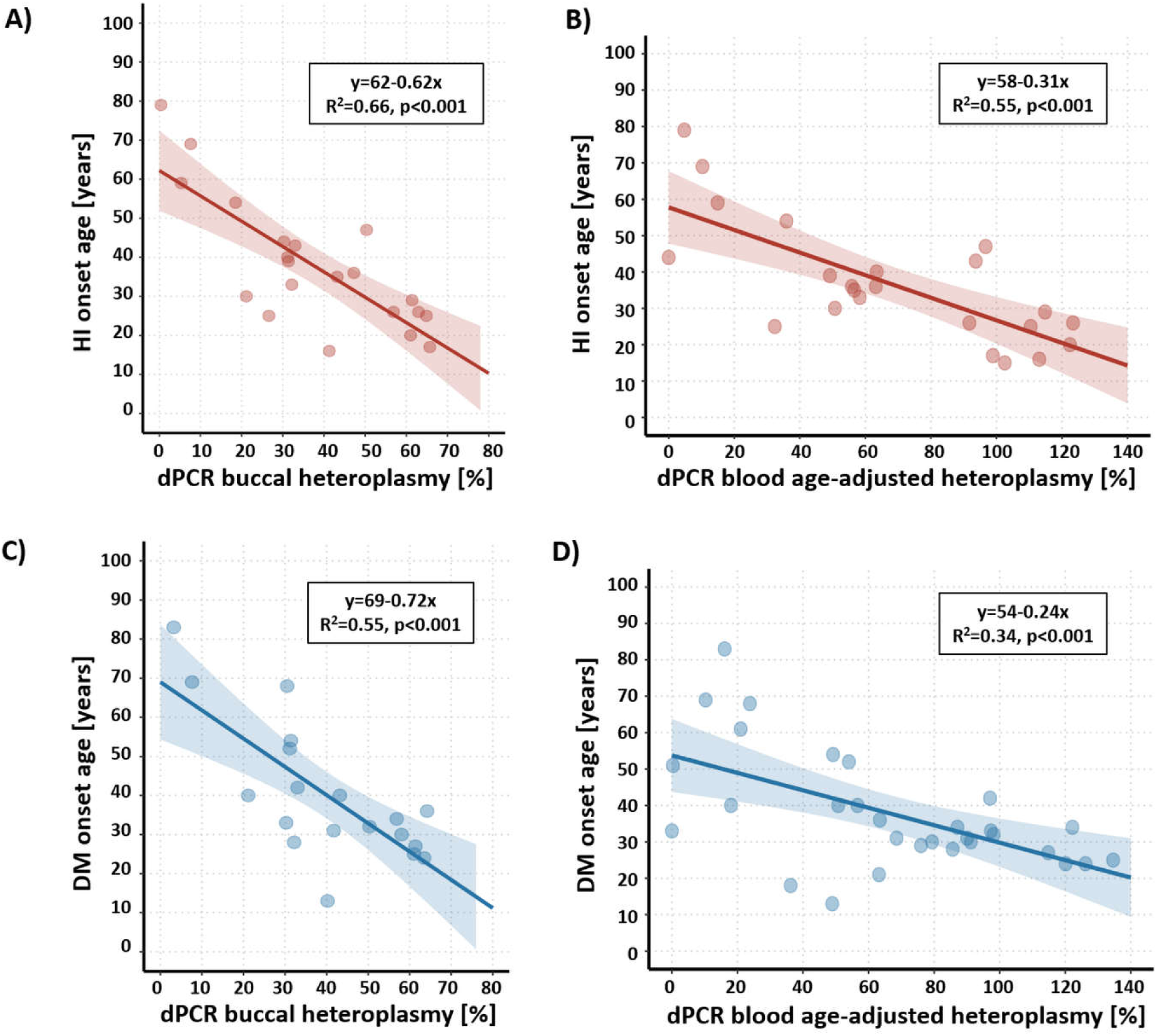
Association of hearing loss onset with (A) buccal heteroplasmy and (B) age-adjusted blood heteroplasmy or association of diabetes mellitus onset with (C) buccal heteroplasmy and (D) age-adjusted blood heteroplasmy in tested individuals carrying the m.3243A>G pathogenic variant. DM – diabetes mellitus; dPCR – digital PCR; HI – hearing impairment.

**Diabetes** has developed in 33 subjects (20 probands and 13 relatives). Originally, it was most frequently diagnosed as T2D (n = 14), although the mean BMI in these T2D patients was 24.02 ± 4.21, and only one of the patients had a BMI higher then 30, which would point to obesity-related T2D. Based on the data provided for 61 subjects carrying the variant, the mean value of BMI was 22.09 ± 5.47. The diabetes type was more rarely labeled as T1D (n = 5), Gestational diabetes mellitus – GDM (n = 2), T1D/T2D overlapping phenotype (n = 1), Impaired glucose tolerance – IGT (n = 1) or directly as MIDD (n = 4) and MELAS (n = 1); data on the originally diagnosed DM type was unavailable in 5 subjects. One more subject had not yet developed diabetes; however, he was newly diagnosed with impaired glucose tolerance based on the Oral Glucose Tolerance Test (OGTT) results in this study. The glycemic control of DM reflected by HbA1c levels did not correlate with blood or buccal heteroplasmy levels (R^2^ ≤ 0.05, *p* > 0.25).

The average age of diabetes onset was 37.62 ± 15.64 years, from 13 to 83 years (Supplementary Figure S2 in the Supplemental Digital Content). Similarly, as for hearing loss, we calculated the linear regression between the diabetes age of onset and heteroplasmy levels. The R^2^ value was higher for the heteroplasmy in buccal mucosa (R^2^ = 0.55, *p* < 0.001) (Figure 2C) than for age-adjusted blood heteroplasmy (R^2^ = 0.34, *p* < 0.01) (Figure 2D), and in a forward stepwise multiple linear regression analysis only buccal mucosa heteroplasmy was significantly associated with the age of diabetes onset (β = -0.653, *p* < 0.001).

Besides typical MIDD or MELAS phenotypes, most of the probands and some of their symptomatic family members also suffered from at least one other symptom or condition known to be associated with the m.3243A>G variant or mitochondrial diseases in general. These findings included: peripheral neuropathy/polyneuropathy, migrainous cephalea, encephalopathy, epilepsy, basal ganglia calcifications, brain cortical atrophy, optic nerve atrophy, macular retinal dystrophy, cataract, vestibular dysfunction, tinnitus, muscular hypotrophy, fatigue, nephropathy, arterial hypertension, WPW syndrome, hypertrophic cardiomyopathy, hypothyroidism, dyslipidemia, pancreatitis, preterm birth and delayed growth (Supplementary Table S1 available upon request from the corresponding author). There was a statistically significant relationship between the age-adjusted blood heteroplasmy levels and the number of related clinical symptoms of the subjects expressed as the phenotype score (ordered ANOVA, Jonckheere-Terpstra test, *p* = 0.0087) (Figure 3A). Typically, the three individuals with MELAS (probands III:1 FAM1, III:1 FAM3 and III:1 FAM13), the most severe phenotype related to the m.3243A>G variant, had the highest age-adjusted blood heteroplasmy, exceeding 100% (126, 134 and 120%, respectively). In contrast, the number of related clinical symptoms did not significantly increase with rising buccal mucosa heteroplasmy levels (ordered ANOVA, Jonckheere-Terpstra test, *p* = 0.529) (Figure 3B). The increasing number of clinical symptoms was also positively associated with the severity of hearing impairment (R^2^ = 0.48, *p* < 0.001) (Figure 3C) and also weakly with glycemic control of DM reflected by HbA1c levels (R^2^ = 0.123, *p* < 0.05).

**Fig. 3.**
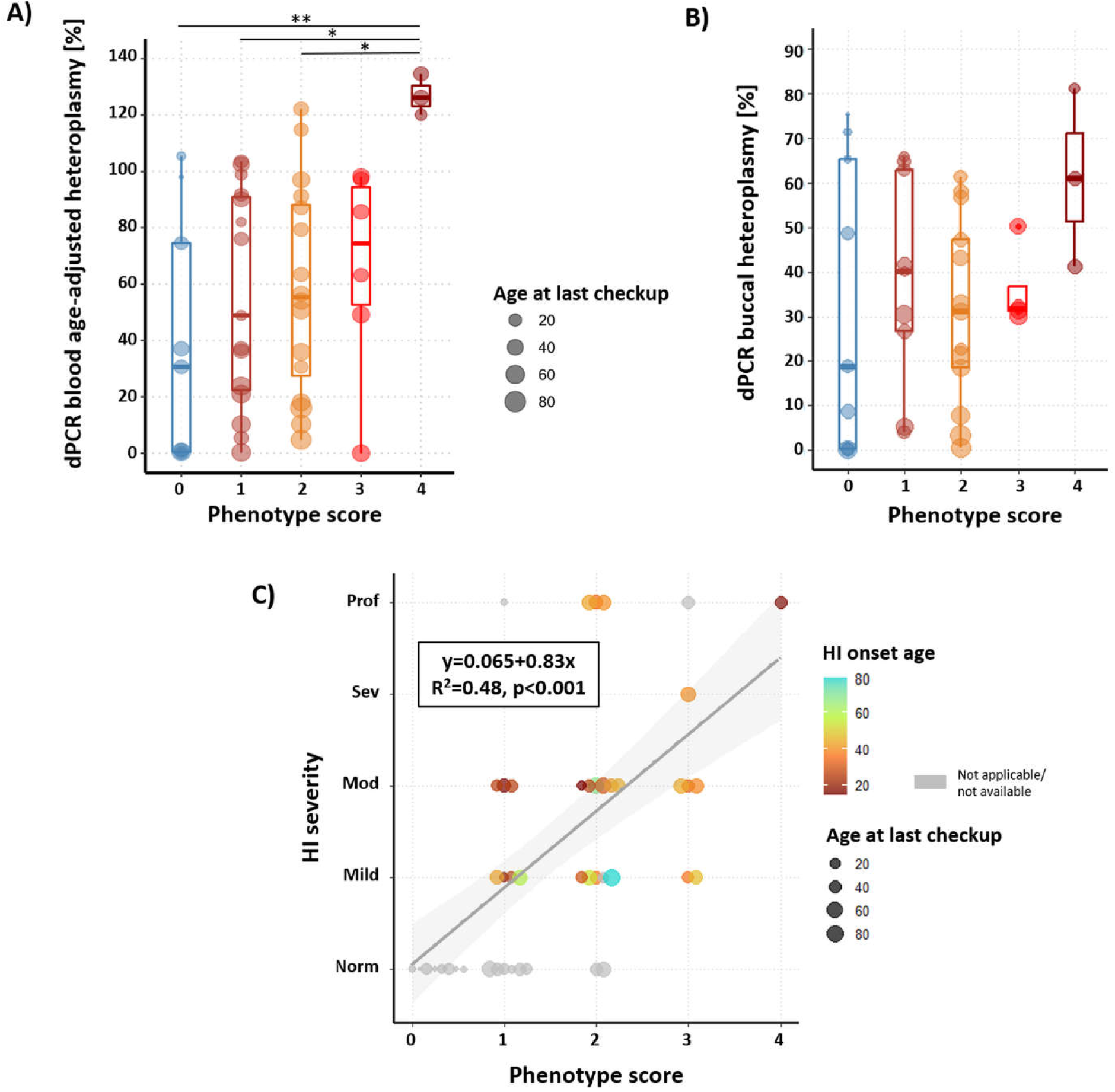
m.3243A>G heteroplasmy levels in the (A) blood or (B) buccal DNA and phenotype score. The point size is relative to the age at last checkup. An open box represents the interquartile range with the line showing the median value. Asterisks indicate significance level of post-hoc Tukey multiple comparison test (* – p<0.05; ** – p<0.01). (C) Association of hearing loss severity with phenotype score (number of symptoms present). The color of the points shows the age at HI onset, while the point size is relative to the age at the last checkup. dPCR – digital PCR; HI – hearing impairment.

Apart from subjects with typical clinical manifestation, we also observed some unexpected findings. Proband III:2, FAM15 (Figure 4A available upon request from the corresponding author) was originally referred for genetic testing due to suspicion of Waardenburg syndrome (WS). She had bilateral hearing loss of moderate degree and iris discoloration resembling heterochromia iridis (Figures 4A and 4B available upon request from the corresponding author). The whole exome sequencing and in-house virtual NGS gene panel (SNHLv9, 438 genes) did not confirm any disease-causing variant in genes known to be associated with WS. Moreover, a detailed investigation of her phenotype showed a mixed hearing loss type and absent acoustic reflexes. The iris discoloration was more precisely classified as a pigmented nevus of the iris in the lower quadrants of the right eye (Figure 4B available upon request from the corresponding author). Her sister III:4, FAM15 (Figure 4A available upon request from the corresponding author) had a light hair strand (Figure 4C available upon request from the corresponding author) and mild low-frequency hearing loss in her left ear. The temporal bone CT scan of the proband showed signs of bilateral small otosclerotic foci in the *fissula ante fenestram*, corresponding to grade II of Veillon classification. Otherwise, the CT scan showed normal ossicular chain and inner ear anatomy (Figure 4D available upon request from the corresponding author). The attempt for exploratory tympanotomy was postponed due to the subsequent pregnancy and maternity leave of this subject. Finally, we performed a sequential stapedotomy on both ears with confirmation of the otosclerosis diagnosis and with excellent results defined by complete closure of the air-bone gaps (≤10 dB). The preoperative and postoperative audiograms are shown in Figure 4A and are available upon request from the corresponding author. Her postoperative hearing thresholds are stable after a three-year follow-up period.

### Long-term cochlear implantation outcomes

Three CI recipients were identified among the subjects genetically tested for the m.3243A>G variant and all belonged to the HL group. Their preoperative audiograms are shown in Supplementary Figure S3 and available upon request from the corresponding author (subjects III:1, FAM3; III:2, FAM12 and II:4, FAM16). All of them were adult female patients implanted unilaterally and had at least five years of experience with CI. The functional hearing outcomes with CI were assessed using a standard battery of tests used for CI recipients, including free field tone audiometry with warble tone, Slovak speech audiometry in noise and categories of auditory performance (CAP) (Table 1). The free field tone audiograms with CI were very similar in all three cases and could be regarded as excellent results (Table 1). In Slovak speech audiometry in noise, all three subjects achieved at least a 50%-word discrimination score, and the CAP score ranging between 6 (understand conversation without lip reading) and 7 (use the telephone with a known speaker). Unfortunately, subject III:1 FAM3 died in his forties (10 years post CI) due to complications associated with the MELAS phenotype.

**Table 1.**
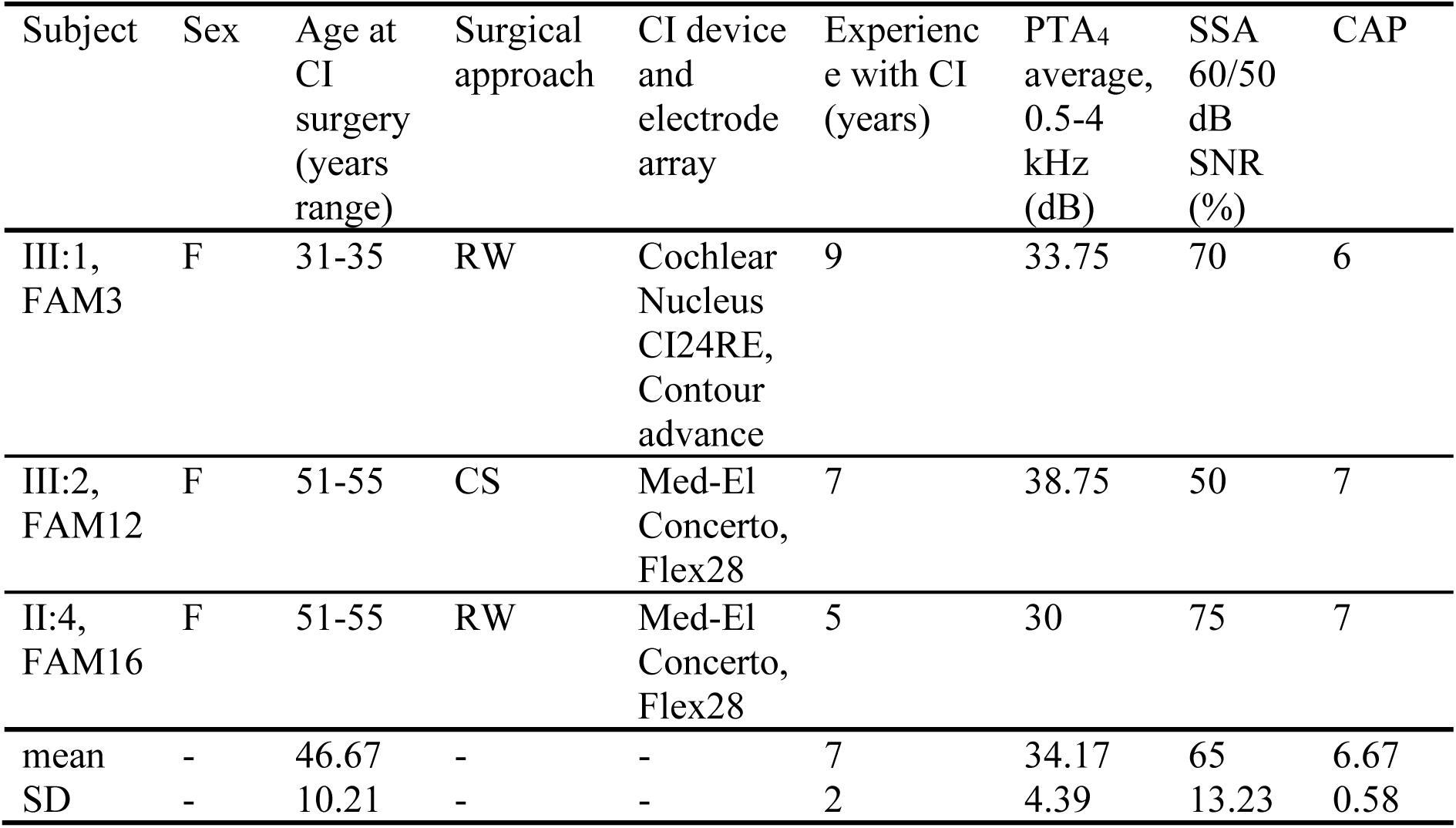
Characterization and selected long-term functional outcomes of cochlear implant users diagnosed with the m.3243A>G variant. CI, cochlear implant; RW, round window; CS, cochleostomy; PTA4, free field tone audiometry thresholds at four frequencies (0.5, 1, 2, and 4 kHz) with CI; SSA, Slovak speech audiometry; SNR, signal-to-noise ratio; CAP, categories of auditory performance.

| Subject | Sex | Age at CI surgery (years range) | Surgical approach | CI device and electrode array | Experience with CI (years) | PTA<sub>4</sub> average, 0.5-4 kHz (dB) | SSA 60/50 dB SNR (%) | CAP |
| --- | --- | --- | --- | --- | --- | --- | --- | --- |
| III:1, FAM3 | F | 31-35 | RW | Cochlear Nucleus CI24RE, Contour advance | 9 | 33.75 | 70 | 6 |
| III:2, FAM12 | F | 51-55 | CS | Med-El Concerto, Flex28 | 7 | 38.75 | 50 | 7 |
| II:4, FAM16 | F | 51-55 | RW | Med-El Concerto, Flex28 | 5 | 30 | 75 | 7 |
| mean | - | 46.67 | - | - | 7 | 34.17 | 65 | 6.67 |
| SD | - | 10.21 | - | - | 2 | 4.39 | 13.23 | 0.58 |

## Discussion

### Prevalence

The prevalence of adult mitochondrial disease, including pathogenic mutations of both mitochondrial and nuclear genomes, was estimated at 1 in 4,300 adults (Gorman et al. 2015). More than half of the known pathogenic variants in mtDNA affect tRNA genes, although tRNA genes contribute to only about 10% of the mitochondrial genome (Moreno-Loshuertos et al. 2011). Among patients with different confirmed mtDNA disorders, the m.3243A>G variant was found to be the most common causal variant, detected in 36% of the cases (Gorman et al. 2015). The prevalence of this variant differs across various studies and is influenced by the geographical population and the selection criteria for subject enrolment. While our data do not provide estimated variant prevalence for the general population in Slovakia, this is estimated to 1:10,000 according to the GnomAD v.4.1.0 population database (Chen et al. 2024). However, the prevalence is influenced by the heteroplasmy threshold used in a given study. Individuals with heteroplasmy <10% are present in GnomAD, but their heteroplasmy levels are not specified. The study of a population cohort of ∼180,000 clinically unselected individuals from the UK Biobank had a threshold at heteroplasmy ≥ 3% in blood and showed much higher prevalence of 1:2167 (Cannon et al. 2024).

Most studies on the prevalence of the m.3243A>G variant have concentrated on patient cohorts with sensorineural hearing loss (SNHL), diabetes or other clinical manifestations linked to MIDD, MELAS and other mitochondrial disease phenotypes. If the main selection criterion is the presence of sensorineural hearing loss, this variant was detected in 0.1–2% of the hearing-impaired cohorts in different geographic populations (Hutchin et al. 2001; Nagata et al. 2001; Montazer Zohour et al. 2012; Tang et al. 2019). In our HL cohort, the prevalence of this variant was similar – 0.79%. At the same time, its prevalence was 3.11% in the subgroup of our patients with postlingual hearing loss, indicating that this mitochondrial variant is a significant causal factor for postlingual hereditary hearing loss in our population.

Concerning another m.3243A>G related symptom – diabetes – it may be misclassified in clinical practice as T1D or T2D depending on the severity of insulin deficiency. It has been reported that approximately 8% of MIDD cases present as T1D with acidosis and ketonuria, but the majority is considered to be T2D at onset. Moreover, retinal and renal manifestations of mitochondrial disease may be confused for diabetic complications (Naing et al. 2014). The mean prevalence of the m.3243A>G variant in European diabetic patients is 0.8%, whereas in Japanese diabetic patients it was almost two-times higher (1.5%) (Murphy et al. 2008). In a Chinese cohort of 1094 hospitalized patients with DM, 0.46% carried this variant, and all of these patients were from the T2D subgroup (Tian et al. 2018). In our diabetes group, the prevalence achieved 0.35%, which aligns with the data reported in the literature. However, we demonstrate that the prevalence of the variant highly depends on the diabetes type. We observed that at least 15% of the identified variant carriers who developed diabetes were previously classified as T1D and 42% as T2D. There was a twelve-times higher prevalence, 1.68%, of the m.3243A>G variant in the subgroup of patients with clinical suspicion of monogenic diabetes than in the cohort of unselected diabetic patients. Mitochondrial diabetes is therefore an important cause of monogenic diabetes in our country. This is in agreement with the findings of Colclough et al., who have shown that this variant was the fourth most common cause of diabetes, present in 8% of their cohort of participants referred for non-syndromic monogenic diabetes testing (Colclough et al. 2022). As the variant was found in all main initially diagnosed diabetes categories (T2D, T1D, GDM, monogenic diabetes), none of the diabetes subtypes can be excluded from a recommendation of genetic testing, although the diagnostic yield and hence the cost-effectiveness differ among the certain diabetes types.

### Phenotypes

In our cohort, 33% of the variant carriers manifested with MIDD, 5% had MELAS and the remaining subjects demonstrated a milder mitochondrial disease phenotype (including 19% with hearing loss without diabetes and/or other manifestations) or were asymptomatic (19%). For carriers of mtDNA disease variants who are still asymptomatic, the term “dormant carrier” has been suggested to distinguish them from healthy carriers of variants with autosomal recessive inheritance (de Laat et al. 2019). A similar proportion of phenotypes was observed by Nesbitt et al. (2013) when analyzing 129 cases with the m.3243A>G mtDNA variant included in the Mitochondrial Disease Patient Cohort Study (UK). In that study, ten percent of patients exhibited the classical MELAS phenotype, 30% had MIDD, 13% had overlap syndromes (MELAS/MIDD, MELAS/chronic progressive external ophthalmoplegia (CPEO), and MIDD/CPEO), 6% demonstrated CPEO and 28% manifested with clinical features not consistent with any of the classical syndromes associated with the m.3243A>G mutation. Isolated sensorineural hearing loss occurred in 3%, and 9% of individuals were asymptomatic. Also, in the study of Fancello et al. (2023), the most common presenting phenotype was MIDD (36.4%) followed by MELAS (30.4%), and the isolated SNHL only occurred in 3.2% of the subjects harboring this variant. The higher proportion of isolated HL or diabetes in our data results from the fact that we screened the cohorts regardless of other than the primary phenotype. This strengthens the view that m.3243A>G testing should also be performed in these cases.

Although the m.3243A>G variant can impact both the inner ear and the nervous system, growing evidence suggests that SNHL caused by mitochondrial pathogenic variants primarily originates in the cochlea (Kullar et al. 2016). Furthermore, we confirmed HL in 57% (36/63) of the subjects with detected mitochondrial variant. In fact, this represents the minimum prevalence of HL in our cohort, because we were unable to obtain precise audiometric data from all subjects originating from the diabetes branch of the study. Hence, at least some cases with milder SNHL could be overlooked. According to earlier published studies, the incidence of sensorineural deafness in patients harboring the mtDNA variant and manifesting DM was estimated to be around 15% (Gerbitz et al. 1995). However, this seems to be highly underestimated. A recent meta-analysis of 161 subjects with mitochondrial diabetes (including 136 cases attributed to the m.3243A>G variant) showed that HL was present in 85.71% of patients with this disease phenotype (Yang et al. 2021). In a review of 272 SNHL cases associated with primary mitochondrial disorders, 68.8% were identified as being associated with the m.3243A>G variant (Fancello et al. 2023).

If we look at the degree of severity of hearing impairment, we see the whole range of hearing impairment levels: mild in 37.5%, moderate to moderately severe in 37.5% and severe to profound in 25%. A similar proportion was described by Fancello et al. (2023), who observed mild HL in 40%, moderate in 34.7% and severe to profound in 25.3%. Generally, SNHL occurs before diabetes (Kade et al. 2020), and the HL in most m.3243A>G carriers is postlingual and progressive. The mean progression rate of hearing loss was 5.5 dB per year based on a retrospective study on 15 Japanese subjects with a mean follow-up period of 12.8 years (Sakata et al. 2022). Our data indicate that HL develops around two years before the onset of diabetes and is postlingual (Supplementary Figure 2). Only one subject manifested prelingual HL (subject III:1, FAM17). However, the early onset of HL in this proband could be associated with risk factors, such as preterm birth, hyperbilirubinemia, bacterial infection and sepsis, as well as hypoxic-ischemic encephalopathy.

We observed significant variability in phenotypes among mutation carriers, along with overlaps with other clinical manifestations, which could potentially lead to misdiagnosis. Most individuals exhibited neuropathy (17.74%), followed by retinopathy (12.9%), encephalopathy (8.06%), myopathy (4.84%), nephropathy (3.23%) and cardiomyopathy (1.61%) (Supplementary Table 1 available upon request from the corresponding author). Similarly, Yang et al. (2021) showed that most MIDD individuals exhibited additional organ manifestations. Central nervous system diseases were found in 29.19%, myopathy in 22.98%, oculopathy in 23.60%, cardiac disease in 23.60% and nephropathy in 13.66% (Yang et al. 2021). Thus, MIDD, similarly to many mitochondrial disorders, is a mitochondrial multiorgan disorder syndrome (MIMODS). Although mitochondrial disorders may manifest in a single organ or tissue at the onset of the disease, other organs are subsequently clinically affected by disease progression (Finsterer & Frank 2017).

### Variability of heteroplasmy levels

Mutated mtDNA can cause mitochondrial diseases when their heteroplasmy levels in the affected tissues and organs exceed a critical threshold. These mutant mtDNAs can be maternally inherited or can arise *de novo*. Rising evidence in animal studies shows that mutant mtDNA levels can vary and change in a non-random fashion across generations and also amongst tissues of an individual. However, the basic cellular and molecular mechanisms of mtDNA heteroplasmy dynamics make it difficult to predict who will inherit or develop mtDNA-associated diseases (Pereira et al. 2021). This trait of uneven heteroplasmy levels and severity of clinical manifestations was also observed in several pedigrees we investigated in this study (Supplementary Figure S3 available upon request from the corresponding author). The difference in heteroplasmy levels between a mother and her offspring may be explained by a severe germ-line mtDNA bottleneck, which may transform a benign (low) frequency in a mother into a disease-causing (high) frequency in her child (Rebolledo-Jaramillo et al. 2014). Interestingly, Uusimaa et al. (2007) have shown that mothers with heteroplasmy greater than 50% tend to have offspring with lower or equal heteroplasmy, whereas the opposite was true for mothers with heteroplasmy less than or equal to 50%. Kopinski et al. (2019) also observed that mutations in the mtDNA cause distinct metabolic and epigenomic changes at different heteroplasmy levels, which could explain the phenotypic variability in mitochondrial disease. According to that study, patients carrying the m.3243A>G variant with heteroplasmy of 20% to 30% commonly present with T1D or T2D or manifest autism. At 50% to 80% heteroplasmy, they may also develop myopathy, cardiomyopathy, lactic acidosis and stroke-like episodes separately or as the complex MELAS syndrome. Subjects with ≥90% heteroplasmy to homoplasmy manifest perinatal lethal diseases, such as Leigh syndrome (Kopinski et al. 2019). Our results confirmed previously published data; we observed a significant correlation between age-adjusted blood heteroplasmy levels and the number of clinical symptoms of the subjects (Figure 3A). All three subjects with MELAS syndrome (probands III:1 FAM1, III:1 FAM3 and III:1 FAM13) had ≥100% age-adjusted blood heteroplasmy.

We used blood leukocytes and buccal mucosa cells to assess the mutation load. Sakata et al. (2022) showed that age-adjusted heteroplasmy levels correlate with the age of onset of hearing loss, and Yang et al. (2021) found a correlation between peripheral blood heteroplasmy levels and age at the onset of diabetes. However, our data indicate that the heteroplasmy of buccal mucosa provides a better prediction of HL onset age (R^2^ = 0.66, *p* < 0.001, Figure 2A) than the age-adjusted blood heteroplasmy (R^2^ = 0.55, *p* < 0.001, Figure 2B). Similarly, only buccal mucosa heteroplasmy was significantly associated with age of diabetes onset (β = -0.653, *p* < 0.001) in a multiple regression. If we look at the onset of the first symptoms in the patients in our study, in most individuals with the MIDD phenotype or isolated hearing loss or diabetes, symptoms appeared in the fourth decade of life. For those with the MELAS phenotype (probands III:1 FAM1, III:1 FAM3 and III:1 FAM13), symptoms manifested as early as in the second decade. The onset of MELAS syndrome is usually between 2 and 31 years, whereas onset after 40 years old is extremely rare (Yang et al. 2022). Our data further indicate that heteroplasmy in buccal mucosa did not significantly predict the severity of HL or diabetes. Conversely, the severity of HL was found to be more closely related to the overall phenotype score (R^2^ = 0.48, *p* < 0.001). Such a correlation was not observed in the case of DM (R^2^ = 0.123, *p* < 0.05).

Our results showed that the interpretation of the data is highly dependent on the tested clinical material. De Laat et al. (2012) found a correlation between the score of the Newcastle Mitochondrial Disease Adult Scale (Schaefer et al. 2006) and heteroplasmy in blood, buccal mucosa and urine, of which buccal mucosa had the best correlation. They concluded that in the general group of m.3243A>G variant carriers the heteroplasmy in buccal saliva is a more effective predictor of clinical outcomes (de Laat et al. 2012). Kullar et al. (2016) attempted to compare the advantages of determining heteroplasmy in blood and urine. Although they found a positive correlation between mean age-corrected pure tone audiogram and urinary heteroplasmy levels, this did not reach statistical significance (r = 0.37, n = 10, *p* = 0.3). No relationship was found between hearing impairment severity and blood heteroplasmy levels (Kullar et al. 2016). However, repeated urine collection within a two-week window in m.3243A>G carriers demonstrated that heteroplasmy levels of urinary epithelial cells are subject to significant day-to-day variation (˃20% margin in one-third of investigated subjects). The interpretation of a correlation between heteroplasmy levels in urine and disease severity may therefore be not reliable (de Laat et al. 2019). Blood is thus the most strongly correlated mutation indicator for assessing disease severity and progression in individuals with the m.3243A>G variant. Blood and urine m.3243A>G heteroplasmy levels are also negatively correlated with age. Blood heteroplasmy declines by ∼2.3%/year, and therefore it is necessary to calculate age-adjusted blood heteroplasmy in routine clinical assessment (Grady et al. 2018). In a previous study of Chae et al. (2020), the mutational load in MELAS patients was reported to inversely correlate with first symptom onset, age at diagnosis of MELAS syndrome and DM. However, the mutational load did not correlate with the clinical severity or progression of DM/IGT. There was no significant difference in insulin resistance or sensitivity indices between the low- and high-mutation load groups (Chae et al. 2020). In our study, data on diabetes therapy was available from 27 subjects with diabetes. Thirteen (48.2%) required insulin treatment, eleven (40.7%) used oral antidiabetics, two (7.4%) were on a diet and one (3.7%) was yet without treatment. However, the majority of subjects on oral antidiabetics (9/11) were administered metformin, which is best to be avoided in subjects with MIDD, as it tends to increase blood lactate levels, leading to lactic acidosis, and trigger neurological manifestations (Schaefer et al. 2013; Tong et al. 2022). In addition, mitophagy, a cellular process that recycles mitochondrial fragments, is known to preferentially eliminate pathogenic heteroplasmic mtDNA mutants. This mechanism may consequently enhance the quality of mtDNA in heteroplasmic mtDNA diseases. It was revealed that metformin also inhibits mitophagy at clinically relevant concentrations (Diot et al. 2015), which might be another reason to avoid metformin.

### m.3243A>G variant and cochlear implantation and stapedotomy outcomes

We identified three CI users harboring the m.3243A>G variant in our hearing loss cohort. All had a clear benefit from rehabilitation with CI and showed excellent functional outcomes, such as the ability to communicate without lip reading or using a telephone. However, literature data on CI outcomes in individuals deafened due to the m.3243A>G variant remains very limited. According to a recent systematic review by Zia et al. (2021), only 11 papers on CI outcomes in mitochondrial hearing loss have been published, and they describe a total of 17 subjects with follow-up ranging between 1 week and 12 months. In total, these studies included 8 subjects with confirmed m.3243A>G variant (Raut et al. 2002; Yasumura et al. 2003; Mancuso et al. 2004; Howes et al. 2008; Yamamoto et al. 2015), with a maximum number of subjects per one paper not exceeding four cases (Yamamoto et al. 2015). Five more published cases were based on the clinical phenotype (MIDD, MELAS), possibly also due to the m.3243A>G variant, but the genetic testing was not performed or its results were not reported (Karkos et al. 2005; Yamamoto et al. 2015; Oliveira et al. 2017). More recently, two more CI cases with the m.3243A>G variant and MELAS syndrome have been reported but again only with short-term postoperative evaluation of up to two years (Crundwell et al. 2022). Thus, we provide the first insight into long-term (≥5 years) CI outcomes in this particular genetic etiology. Nevertheless, all data published to date indicate that the vast majority of patients with MIDD/MELAS syndrome and the m.3243A>G variant may achieve excellent or near-excellent functional outcomes with CI, and based on our results, these outcomes are stable over a longer period. However, it is important to consider the potential risks of general anesthesia and cognitive decline in patients with advanced-stage MELAS (Chinnery et al. 2000; Crundwell et al. 2022). In addition, MRI follow-up in patients with encephalopathy and stroke-like episodes may be severely limited due to metallic artifacts from the implant.

Further, through this screening study, we identified more subjects (III:1 FAM1, III:1 FAM11, III:1 FAM17) who could potentially benefit from CI but were either unaware of this hearing rehabilitation option or had not yet been referred to the CI center by their caretaking physicians. The presence of other clinical features seen in FAM15 (pigmentary disorders, otosclerosis) is most likely only a coincidence with the m.3243A>G variant. Moreover, this family did not show any of the typical clinical manifestations associated with the m.3243A>G variant, such as diabetes or pure sensorineural hearing loss. The only clinical feature commonly associated with this mitochondrial variant was the low BMI (18.4 at the age of 31-35 years) of the proband. To date, no causal relationship has been established between otosclerosis and pathogenic mitochondrial variants despite extensive research on hereditary hearing loss. Similarly, to the best of our knowledge, no case of otosclerosis and the m.3243A>G mtDNA variant diagnosed simultaneously has been previously reported in the scientific literature. Thus, based on current knowledge, we consider these two findings as two separate diagnoses. However, they may have a synergic effect on the progression of hearing loss in the future. Therefore, long-term audiometric follow-up of the subject will be necessary to record any changes in the bone conduction thresholds. The presented case is also the first reported case of successful stapedotomy in the presence of a disease-causing mtDNA variant. Here we provide evidence that this surgical procedure in which the inner ear is opened by creating a small fenestra in the stapedial footplate may be safe and efficient in patients with impaired mitochondrial metabolism of the inner ear. We believe that the current hearing loss phenotype in proband III:2 FAM15 is most probably determined only by the otosclerosis, but due to patient’s still relatively young age, the m.3243A>G variant may become manifest in the future. If that were the case, it would confirm the hypothesis that the presence of additional genetic factors may explain the variable degree of hearing loss progression in patients with otosclerosis.

Similarly, there are no available literature data on WS-like pigmentary changes associated with the m.3243A>G variant. In contrast, in 15–70% of WS cases depending on the WS type, the etiology remains unexplained at the molecular level (Pingault et al. 2010). Cases of WS harboring autosomal dominant variants often manifest by incomplete phenotypes, including individuals with pigmentary disorder but normal hearing, as we reported in our previous study (Varga et al. 2021). However, the original suspicion of WS in the family presented in this paper was a misdiagnosis based on a combination of pigmentary disorder and hearing loss, which was excluded by detailed clinical evaluation and molecular-genetic investigation.

It is also important to underline the limitations of this study. They mostly result from different inclusion criteria and recruitment strategies in the three patient cohorts. As a consequence, the availability of certain biological material (buccal swabs to determine buccal mucosa heteroplasmy) or different extent of clinical data obtained (e.g., less detailed data on HL in the diabetic group) may have an impact on the quality of the data provided.

## Conclusion

We achieved a higher diagnostic yield in the deafness and monogenic diabetes groups than in the unselected diabetes group. This is in agreement with the fact that these two groups were formed using criteria favoring inherited forms. The severity of mitochondrial disease related to the m.3243A>G variant is highly variable and can range from severe multisystemic involvement through isolated symptoms to asymptomatic young “dormant” or very low heteroplasmy carriers. The implementation of **strict inclusion criteria** requiring the presence of both diabetes and hearing loss in our study would lead to about a 54% lower detection rate due to different or incomplete phenotype manifestation. Regarding the **predictability** of disease progression, heteroplasmy levels from buccal swabs showed a better correlation with the age of onset of both hearing loss and diabetes than the age-adjusted blood heteroplasmy. On the other hand, age-adjusted blood heteroplasmy was associated with overall disease severity (i.e., with a higher number of clinical symptoms). Interestingly, the severity of hearing loss could not be predicted by heteroplasmy in either of the two tissue types, but it was related to the overall severity of disease. Although causal **treatment** for patients carrying the mitochondrial m.3243A>G variant is not yet available, the variability in symptom onset and clinical manifestation underlines the need for individualized care and the importance of early diagnosis. From this perspective, we further confirm that even cochlear **implantation and stapedotomy** are safe and efficient options for long-term hearing restoration and rehabilitation in m.3243A>G carriers.

The manuscript was edited according to the requirements of medRxiv, and the detailed clinical data are available upon request from the corresponding author.

## Data Availability

Clinical data produced in the present study are available upon reasonable request to the corresponding author.

## Acknowledgments

We would like to thank our mentor, prof. Iwar Klimes, who initiated research on mitochondrial diabetes in Slovakia. Sadly, he passed away before this study could be completed. We are also grateful to all collaborating physicians and patients who participated in this study. This work was financially supported by the research grants APVV-20-0236, VEGA 1/0572/21, VEGA 2/0131/21, ITMS 26240120038 and APVV-17-0296.

## Authors’ contribution

Conceptualization of the study: D.G., L.V., M.P.; writing of the manuscript: L.V., S.B, M. Sko.; molecular-genetic analyses performed: V.R., M. Skl., K.C.; clinical evaluation performed: L.V., J.S., S.B., D.U., Z.K.; statistical analyses performed: M.Sko., L.V.; graphic material prepared: S.B., M. Sko., L.V. All authors discussed the results and reviewed the manuscript.

## Notes

**Financial disclosures/conflicts of interest**: There are no conflicts of interest, financial, or otherwise.

### Competing Interest Statement

The authors have declared no competing interest.

### Author Declarations

The study was conducted in accordance with the Declaration of Helsinki and the Guidelines for Good Pharmacoepidemiology Practice. The study was reviewed and approved by institutional review board/ethics committee - Ethics Committees of the University Hospital in Bratislava (registered with clinicaltrials.gov as NCT02232503), the Ethics Committees of the University Hospital in Bratislava (No. 33/26102010, PI: Milan Profant), the National Institute of Childrens Diseases in Bratislava (No. 14/06/2005, PI: Lubomir Barak) and the National Institute of Endocrinology and Diabetes in Lubochna (No. 19/03/2007, PI: Jozef Michalek), before the study began.

