## Supplementary Figure S1 for "Screening for Maternally Inherited Diabetes and Deafness in Large Cohorts of Hearing Impaired and Diabetic Patients"

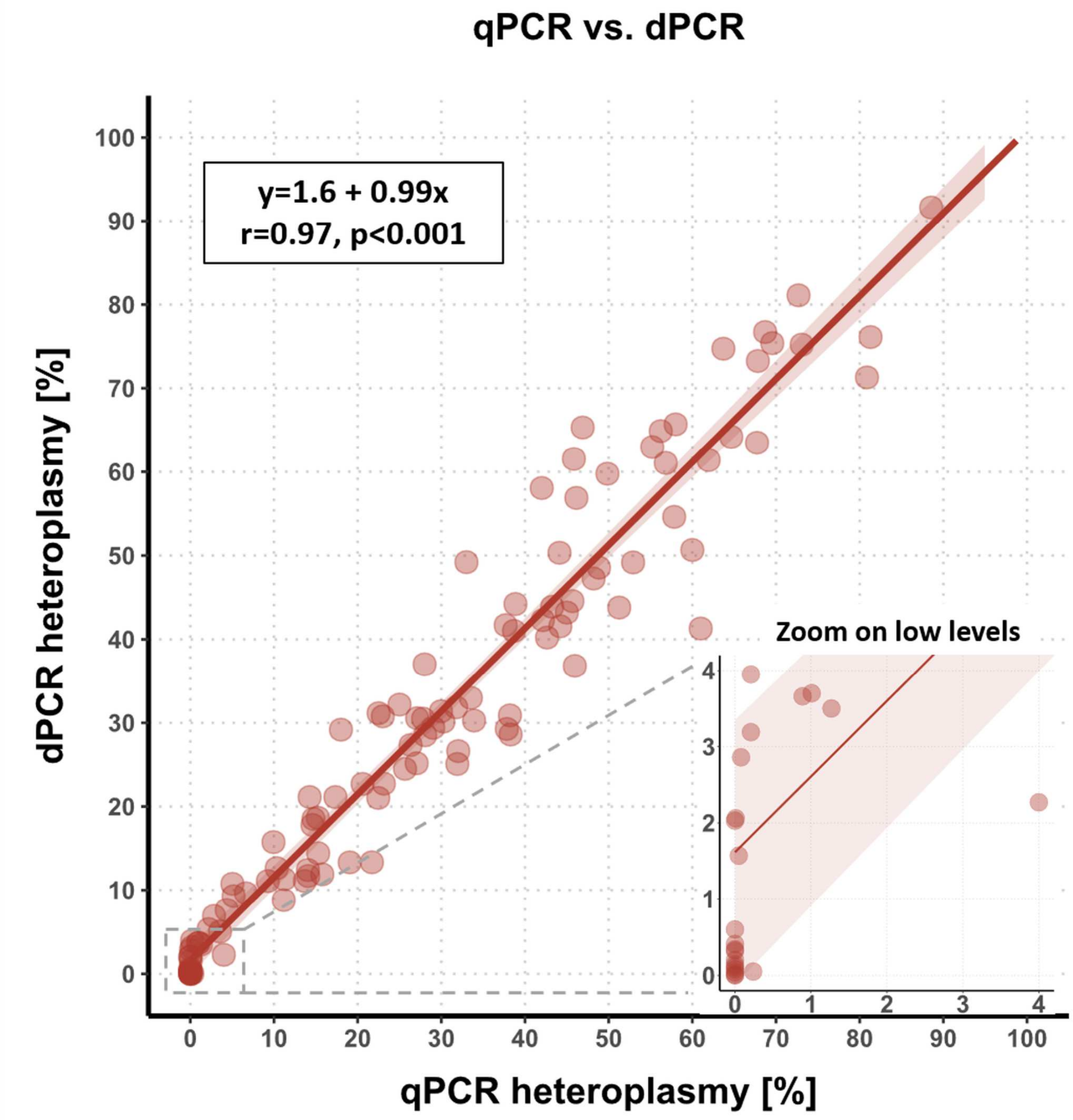


**Supplementary figure 1:** Correlation of heteroplasmy levels obtained by two methods – qPCR and dPCR. The zoomed-in window shows higher sensitivity of dPCR in lower heteroplasmy levels. r – correlation coefficient counted using Stat_cor function (ggpubr R package).
