## Supplementary Figure S2 for "Screening for Maternally Inherited Diabetes and Deafness in Large Cohorts of Hearing Impaired and Diabetic Patients"

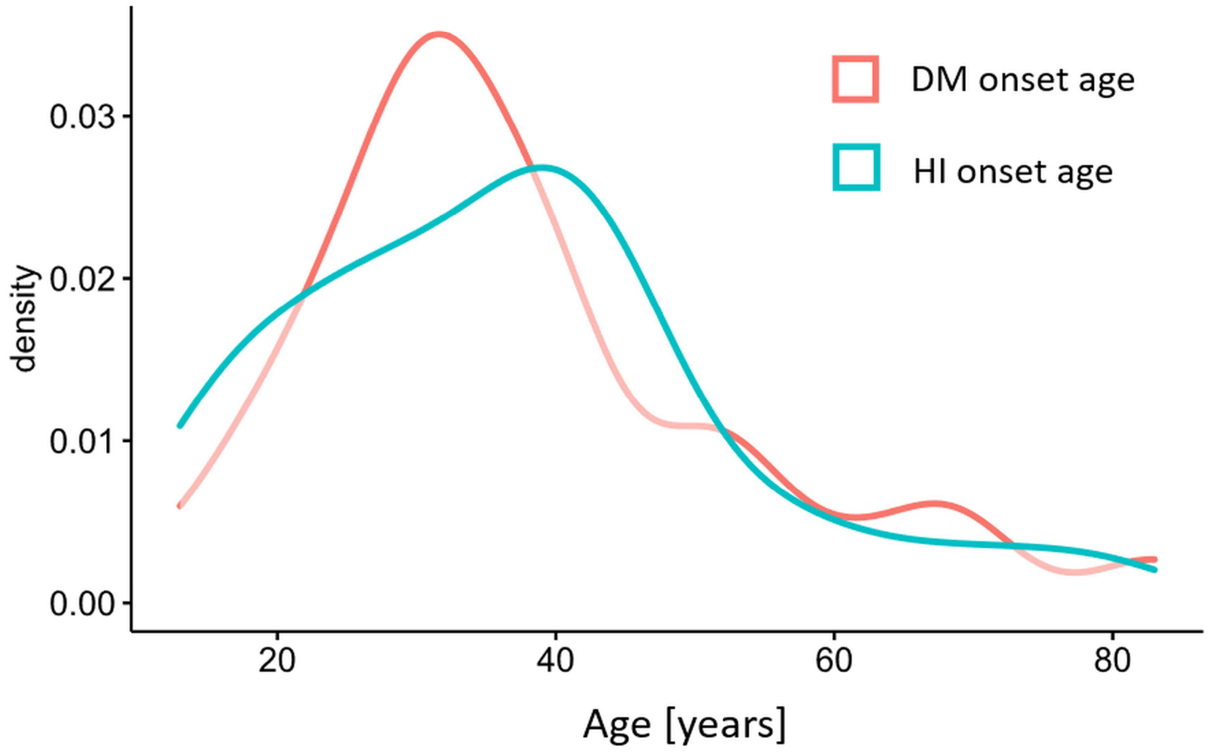


**Supplementary figure 2:** Age of onset of diabetes mellitus (red) and hearing impairment (blue) distribution in tested cohort.
